# Cognitive screening instruments for dementia: comparing metrics of test limitation

**DOI:** 10.1101/2020.10.29.20222109

**Authors:** Andrew J Larner

## Abstract

Cognitive screening instruments (CSIs) for dementia and mild cognitive impairment are usually characterised in terms of measures of discrimination such as sensitivity, specificity, and likelihood ratios. However, CSIs also have limitations. Several metrics exist which may be used to denote test limitations but they are seldom examined. Data from several pragmatic test accuracy studies of CSIs were interrogated to calculate various measures of limitation, namely: misclassification rate; net harm to net benefit ratio; and the likelihood to be diagnosed or misdiagnosed. Intra- and inter-test performance for measures of discrimination and limitation were compared. The study found that some tests with very high sensitivity but low specificity for dementia fared poorly on measures of limitation, with high misclassification rates, low net harm to net benefit ratios, and low likelihoods to be diagnosed or misdiagnosed; some had likelihoods favouring misdiagnosis over diagnosis. Tests with a better balance of sensitivity and specificity fared better on measures of limitation. When choosing which CSIs to administer, measures of test limitation should be considered as well as measures of test discrimination. Although high test sensitivity may be desirable to avoid false negatives, false positives also have a cost. Identification of tests having high misclassification rate, low net harm to net benefit ratio, and low likelihood to be diagnosed or misdiagnosed, may have implications for their use in clinical practice.

## Introduction

Like all screening and diagnostic tests, cognitive screening instruments (CSIs) are usually characterised in terms of the conditional probabilities of sensitivity (Sens) and specificity (Spec), where Sens (or true positive rate, TPR) is the correct identification of those with dementia or cognitive impairment, and Spec (or true negative rate, TNR) is the correct exclusion of those without disease (see Box 1 for definitions of metrics discussed in this paper, their formulae and score ranges). Information from both Sens and Spec may be combined in metrics such as the Youden index (Y) and positive and negative likelihood ratios (LR+, LR-), of which the latter may be qualitatively classified as causing slight, moderate, large, or very large change in probability of disease or its absence.^1^ Sens and Spec are suggested keywords for reports of diagnostic test accuracy studies in dementia (STARDdem)^2^ and LRs were used as the basis for recommendations made by the UK National Institute for Health and Care Excellence for tests suitable for dementia.^3^ Systematic reviews and meta-analyses of CSIs, for example those produced by the Cochrane Dementia and Cognitive Improvement Group,^4^ typically quote summary test Sens, Spec, and LRs.

Like all screening and diagnostic tests, CSIs are not perfect. They have shortcomings, inadequacies, or failures, which may be termed “limitations”. Tests have potential harms (misdiagnosis) as well as benefits (correct diagnosis). The limitations comprise failure to identify dementia or cognitive impairment when it is in fact present and identifying these states when they are in fact absent. These rates, respectively false negative (FNR) and false positive (FPR), are implicit in the measures of Sens and Spec since, by the principle of summation, they are their complements or negations (FNR = 1 – Sens; FPR = 1 – Spec). Other metrics of test limitation include inaccuracy (Inacc; also sometimes known as fraction incorrect, or error rate) and error odds ratio (EOR), although these measures are seldom used in clinical practice.

Other metrics of test limitation, which, like all those already mentioned, may be derived from the 2×2 contingency table of diagnostic test accuracy studies, form the subject of the current study. These are: the misclassification rate; the net harm to net benefit ratio; and the likelihood to be diagnosed or misdiagnosed.

The sum of FNR and FPR is used here to define the misclassification rate (MR), following the usage of Perkins and Schisterman.^5^ (Confusingly this term has also been sometimes used interchangeably with Inacc.) Minimization of MR is used in some of the methods for setting a test threshold from inspection of the receiver operating characteristic (ROC) curve of a test accuracy study.

The ratio of net harm to net benefit (H/B) may be defined as the harm (H) of treating a person without disease (i.e. false positive) to the net benefit (B) of treating a person with disease (i.e. true positive), the latter term equating to the harm of a false negative result.^6^ The H/B ratio may be calculated from the Bayes’ equation as the product of the pre-test odds of disease and the positive likelihood ratio at the specified test cut-off (which is equivalent to the slope of the ROC curve, TPR/FPR, at that point) and hence is equivalent to the post-test odds.^7^ A higher H/B ratio means the test is less likely to miss cases, and hence less likely to incur the harms of false negatives, and hence a higher H/B ratio is deemed better (note that this scoring of H/B ratio may seem counterintuitive if one thinks solely of “harms” and “benefits”, hence the important qualification of “net”).

More recently, another metric attempting to denote test limitation has been introduced: the likelihood to be diagnosed or misdiagnosed (LDM).^8,9^ LDM is based on “number needed” metrics which are generally deemed to be more intuitive and hence applicable for both clinicians and patients than Sens and Spec. One form of LDM is given by the ratio of the number needed to misdiagnose,^10^ which is the inverse of Inacc, to the number needed to diagnose, which is the inverse of Youden index. Hence LDM may also be conceptualised as a ratio of harms (misdiagnosis) and benefits (diagnosis). LDM ranges from 0 to infinity but, as for likelihood ratios, has an inflection point at 1 such that LDM<1 indicates a test in which misdiagnosis is overall more likely than diagnosis, and LDM>1 indicates a test in which diagnosis is overall more likely than misdiagnosis, and hence LDM>>1 is desirable and LDM = ∞ is the perfect diagnostic test (where Sens = Spec = Y =1, and Inacc = 0).

The purpose of this study was to compare these three indices of test limitation (MR, H/B ratio, LDM) for several brief CSIs in common clinical usage for dementia diagnosis, namely the Mini-Mental State Examination (MMSE),^11^ the Montreal Cognitive Assessment (MoCA),^12^ the Mini-Addenbrooke’s Cognitive Examination (MACE),^13^ the Six-item Cognitive Impairment Test (6CIT),^14^ the informant Ascertain Dementia 8 (iAD8),^15^ and the Test Your Memory (TYM) test,^16^ as well as for a more recently described instrument, Free-Cog.

## Methods

Data from previously undertaken and reported pragmatic prospective test accuracy studies^17^ were used (Table 1), each undertaken using a standardised methodology in a dedicated cognitive disorders clinic located in a regional neuroscience centre. In all studies subjects gave informed consent and study protocol was approved by the institute’s committee on human research (Walton Centre for Neurology and Neurosurgery Approval: N 310).

**Table 1:** Study demographics.

| **CSI** | **N** | **Age,**<br>**median**<br>**(years)** | **Gender**<br>**(F:M; %F)** | **Test threshold**<br>**for dementia** | **Pre-test**<br>**odds** |
| --- | --- | --- | --- | --- | --- |
| MMSE | 244 | 60 | 117:127; 48 | <26/30 | 0.22 |
| MoCA | 260 | 59 | 118:142; 45 | <26/30 | 0.21 |
| MACE | 755 | 60 | 352:403; 47 | ≤25/30 | 0.18 |
| 6CIT | 245 | 59 | 121:124; 49 | >9/28 | 0.25 |
| AD8 | 212 | 64.5 | 106:106; 50 | ≥2/8 | 0.43 |
| TYM | 224 | 62 | 94:130; 42 | ≤42/50 | 0.54 |
| Free-Cog | 141 | 62 | 61:80; 43 | ≤22/30 | 0.12 |

Abbreviations: CSI = Cognitive screening instrument; MCI = mild cognitive impairment; TP = true positive; FN = false negative; MMSE = Mini-Mental State Examination; MoCA = Montreal Cognitive Assessment; MACE = Mini-Addenbrooke's Cognitive Examination; 6CIT = Six-item Cognitive Impairment Test; AD8 = Ascertain Dementia 8; TYM = Test Your Memory

In each study, criterion diagnosis of dementia followed standard diagnostic criteria (DSM-IV) and was made independent of scores on index CSIs to avoid review bias. Cross classification of criterion diagnosis with index test result, dichotomised by test cut-off, in a standard 2×2 contingency table allowed all cases to be classified as true positive (TP), false positive (FP), false negative (FN) and true negative (TN). Where possible, test cut-offs documented in the respective index studies were used to avoid bias. All studies followed either the STAndards for the Reporting of Diagnostic accuracy studies (STARD)^18^ or the derived guidelines specific for dementia studies, STARDdem,^2^ dependent on the exact date at which each test accuracy study was undertaken.

## Results

Examining measures of test discrimination (Table 2), many were highly sensitive (MoCA, Free-Cog, MACE, AD8) but had low specificity (MoCA, MACE, AD8). Positive likelihood ratios were qualitatively either slight (MoCA, MACE, AD8, TYM) or moderate (6CIT, Free-Cog, MMSE), none achieving the large or very large classification.

**Table 2:** Comparing metrics of test discrimination for CSIs for diagnosis of dementia vs no dementia (with ranking in brackets)

| **CSI** | **Sens** | **Spec** | **LR+ and qualitative classification** |
| --- | --- | --- | --- |
| MMSE | 0.86 (7) | 0.64 (3) | 2.37 = moderate (3) |
| MoCA | 1.00 (1=) | 0.31 (6) | 1.46 = slight (5) |
| MACE | 0.99 (3) | 0.32 (5) | 1.45 = slight (6) |
| 6CIT | 0.88 (6) | 0.78 (1) | 4.00 = moderate (1) |
| AD8 | 0.98 (4) | 0.10 (7) | 1.09 = slight (7) |
| TYM | 0.95 (5) | 0.45 (4) | 1.73 = slight (4) |
| Free-Cog | 1.00 (1=) | 0.67 (2) | 3.07 = moderate (2) |

Abbreviations: CSI = Cognitive screening instrument; MMSE = Mini-Mental State Examination; MoCA = Montreal Cognitive Assessment; MACE = Mini-Addenbrooke's Cognitive Examination; 6CIT = Six-item Cognitive Impairment Test; AD8 = Ascertain Dementia 8; TYM = Test Your Memory; Sens = Sensitivity; Spec = Specificity; LR+ = Positive likelihood ratio

Examining measures of test limitation (Table 3), few achieved a misclassification rate of ≤0.5 (Free-Cog, 6CIT, MMSE). Only one test (6CIT) achieved H/B ratio of 1. LDM values of <1 (likelihood of misdiagnosis greater than correct diagnosis) were recorded for some tests (MoCA, MACE, AD8). Of note, the tests with high sensitivity but low specificity generally fared worse on these metrics examining test limitation, whilst those with a better balance of Sens and Spec (reflected in the higher LR+s) did better. This was also evident in the overall ranking of CSIs by outcome of the examined measures of discrimination and limitation (Table 4).

**Table 3:** Comparing metrics of test limitation for CSIs for diagnosis of dementia vs no dementia (with ranking in brackets)

| **CSI** | **MR**<br>**(lower**<br>**better)** | **H/B**<br>**(higher**<br>**better)** | **LDM**<br>**(higher**<br>**better)** |
| --- | --- | --- | --- |
| MMSE | 0.50 (3) | 0.52 (3) | 1.56 (3) |
| MoCA | 0.69 (5=) | 0.30 (6) | 0.54 (5) |
| MACE | 0.69 (5=) | 0.26 (7) | 0.53 (6) |
| 6CIT | 0.34 (2) | 1.00 (1) | 3.30 (1) |
| AD8 | 0.92 (7) | 0.47 (4) | 0.12 (7) |
| TYM | 0.60 (4) | 0.93 (2) | 1.08 (4) |
| Free-Cog | 0.33 (1) | 0.38 (5) | 2.31 (2) |

Abbreviations: CSI = Cognitive screening instrument; MMSE = Mini-Mental State Examination; MoCA = Montreal Cognitive Assessment; MACE = Mini-Addenbrooke's Cognitive Examination; 6CIT = Six-item Cognitive Impairment Test; AD8 = Ascertain Dementia 8; TYM = Test Your Memory; MR = Misclassification rate; H/B = Net harm/net benefit ratio; LDM = Likelihood to be diagnosed or misdiagnosed

**Table 4:** Ranking of CSIs by outcome measures of test discrimination and limitation.

| Rank | Measures of test discrimination |  |  | Measures of test limitation |  |  |
| --- | --- | --- | --- | --- | --- | --- |
|  | Sens | Spec | LR+ | MR | H/B | LDM |
| 1 | MoCA,<br>Free-Cog | 6CIT | 6CIT | Free-Cog | 6CIT | 6CIT |
| 2 | - | Free-Cog | Free-Cog | 6CIT | TYM | Free-Cog |
| 3 | MACE | MMSE | MMSE | MMSE | MMSE | MMSE |
| 4 | AD8 | TYM | TYM | TYM | AD8 | TYM |
| 5 | TYM | MACE | MoCA | MACE,<br>MoCA | Free-Cog | MoCA |
| 6 | 6CIT | MoCA | MACE | - | MoCA | MACE |
| 7 | MMSE | AD8 | AD8 | AD8 | MACE | AD8 |

Abbreviations: CSI = Cognitive screening instrument; MMSE = Mini-Mental State Examination; MoCA = Montreal Cognitive Assessment; MACE = Mini-Addenbrooke's Cognitive Examination; 6CIT = Six-item Cognitive Impairment Test; AD8 = Ascertain Dementia 8; TYM = Test Your Memory; Sens = Sensitivity; Spec = Specificity; LR+ = Positive likelihood ratio; MR = Misclassification rate; H/B = Net harm/net benefit ratio; LDM = Likelihood to be diagnosed or misdiagnosed

## Discussion

This study has various shortcomings. The findings are of course dependent upon the diagnostic test accuracy studies upon which they are based. These index studies obviously have limitations, for example they were undertaken in different patient populations, albeit all seen in the same clinic and operating the same diagnostic criteria for dementia, and hence may not necessarily be generalizable. Nevertheless the findings suggest significant limitations for many of the CSIs in common usage. The findings might be corroborated by undertaking similar analyses with data reported in systematic reviews of these CSIs where available.

The measures of limitation examined here are seldom used in clinical practice, may be unfamiliar to clinicians, and have no exact ranges. Other methods of assessing test effectiveness and limitation are also available. The metrics examined here do not address utilities^7^ or cost ratios.^19^

For MR and the H/B ratio, lower or higher values respectively may be better, but precisely how high or how low is most desirable or optimal has not been defined. LDM values have clearer implications around the inflection point of 1. The influence of disease prevalence on MR is unknown, although as it is based (like Sens, Spec, FPR, and FNR) on strict columnar ratios from the 2×2 contingency table it is notionally uninfluenced by the base rate, although it is well recognised that these measures are affected by the heterogeneity (spectrum bias) of clinical populations.^20^ Another formulation of LDM, with the denominator based on predictive values, takes account of disease prevalence.^8,9,17^

Whilst clinicians may be content to use highly sensitive tests, accepting false positives as a reasonable trade-off to ensure no cases are missed (i.e. low false negative rate), metrics of limitation highlight the potential shortcomings of such tests, and emphasize the need to find better tests. Patients undergoing testing may also want to have easily assimilated information on how well the test performs (a false positive diagnosis may have more significance for a patient than for a clinician) as well as its potential risks. Newer biomarker tests of dementia disorders could be subjected to similar analyses of test limitation.

## Data Availability

Data available on any reasonable request

---

**Box 1:**
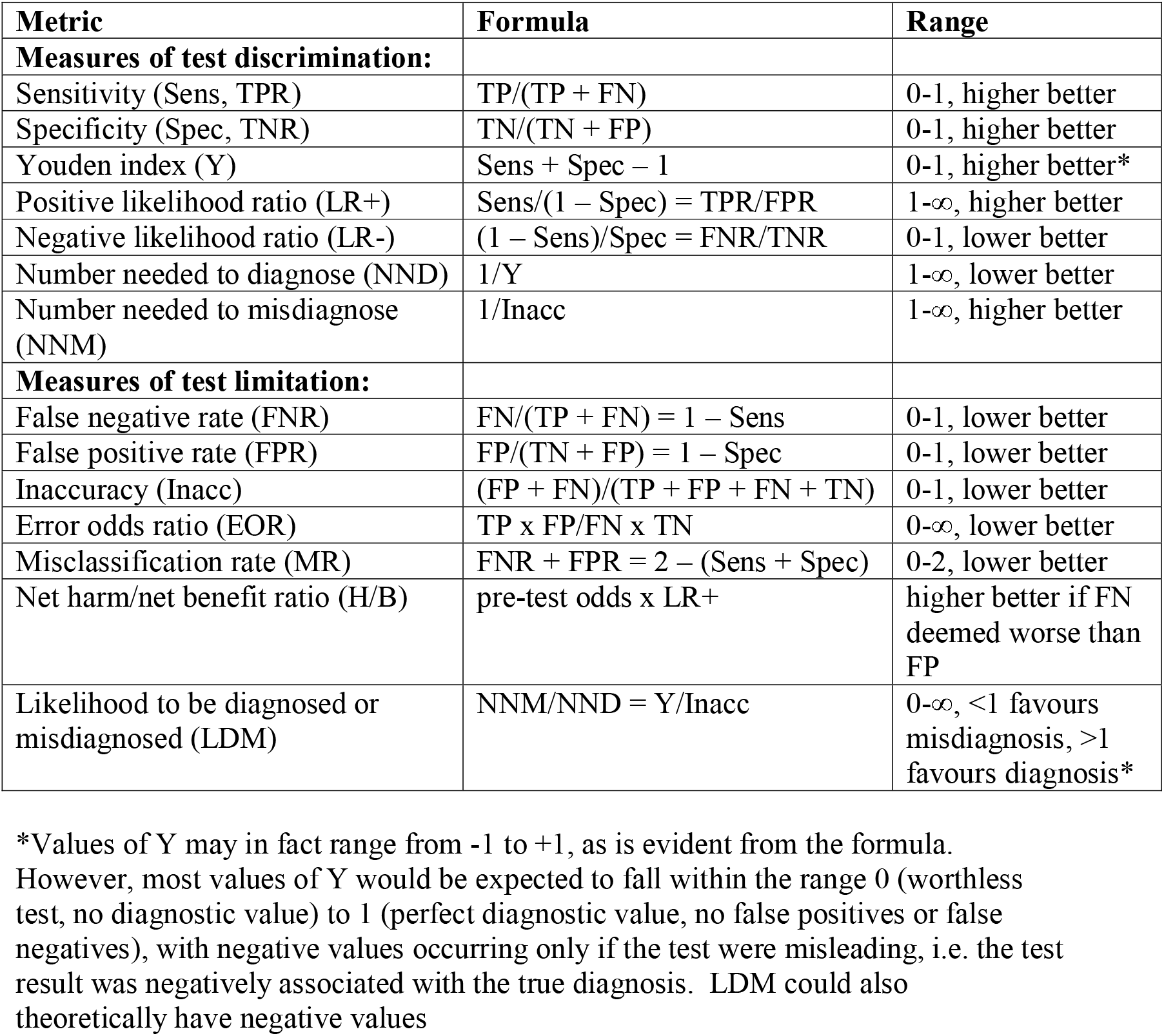
Metrics, formulae, and ranges for measures of test effectiveness and limitation.

